# Automated Thyroid Ultrasound Analysis: Hashimoto’s Thyroiditis

**DOI:** 10.1101/2024.04.24.24306100

**Authors:** Luís Jesuíno de Oliveira Andrade, Gabriela Correia Matos de Oliveira, Luísa Correia Matos de Oliveira, Luís Matos de Oliveira

## Abstract

**Introduction:** Thyroid ultrasound provides valuable insights for thyroid disorders but is hampered by subjectivity. Automated analysis utilizing large datasets holds immense promise for objective and standardized assessment in screening, thyroid nodule classification, and treatment monitoring. However, there remains a significant gap in the development of applications for the automated analysis of Hashimoto’s thyroiditis (HT) using ultrasound.

**Objective:** To develop an automated thyroid ultrasound analysis (ATUS) algorithm using the C# programming language to detect and quantify ultrasonographic characteristics associated with HT.

**Materials and Methods:** This study describes the development and evaluation of an ATUS algorithm using C#. The algorithm extracte relevant features (texture, vascularization, echogenicity) from preprocessed ultrasound images and utilizes machine learning techniques to classify them as “normal” or indicative of HT. The model is trained and validated on a comprehensive dataset, with performance assessed through metrics like accuracy, sensitivity, and specificity. The findings highlight the potential for this C#-based ATUS algorithm to offer objective and standardized assessment for HT diagnosis.

**Results:** The program preprocesses images (grayscale conversion, normalization, etc.), segments the thyroid region, extracts features (texture, echogenicity), and utilizes a pre-trained model for classification (“normal” or “suspected Hashimoto’s thyroiditis”). Using a sample image, the program successfully preprocessed, segmented, and extracted features. The predicted classification (“suspected HT”) with high probability (0.92) aligns with the pre-established diagnosis, suggesting potential for objective HT assessment.

**Conclusion:** C#-based ATUS algorithm successfully detects and quantifies Hashimoto’s thyroiditis features, showcasing the potential of advanced programming in medical image analysis.

## INTRODUCTION

Thyroid ultrasound (US) is a widely used imaging modality for the assessment of thyroid gland structure and function. It is a non-invasive, readily available, and relatively inexpensive technique that provides valuable information for the diagnosis and management of thyroid disorders.^1^ However, the interpretation of thyroid US images can be subjective and operator-dependent, leading to potential variability in diagnostic accuracy.

Manual thyroid US analysis relies heavily on the expertise and experience of the sonographer. This can lead to inconsistencies in image interpretation and reporting, particularly among less experienced practitioners.^2^ Additionally, the subjective nature of manual assessment can be influenced by factors such as fatigue, visual acuity, and individual interpretation biases.^3^

Automated thyroid US analysis (ATUS) offers the potential to address the limitations of manual interpretation by providing objective and standardized assessments.^4^ ATUS algorithms can be trained on large datasets of thyroid US images with corresponding clinical data to identify and quantify subtle US features associated with various thyroid disorders.^5^

ATUS has the potential to be applied in various clinical settings, including: Screening for thyroid disorders: ATUS could be used to screen asymptomatic individuals for thyroid abnormalities, potentially leading to early detection and intervention;^6^ Diagnosis and classification of thyroid nodules: ATUS could assist in the diagnosis and classification of thyroid nodules, helping to differentiate between benign and malignant lesions;^7^ Monitoring of thyroid disorders: ATUS could be used to monitor the response to treatment for thyroid disorders, providing objective measures of disease progression or regression.^8^

Research in ATUS has made significant progress in recent years. Several studies have demonstrated the potential of ATUS algorithms to accurately differentiate between normal and abnormal thyroid tissue, classify thyroid nodules, and monitor treatment response.^9^ However, further validation and refinement are needed before ATUS can be widely adopted in clinical practice.

The objective of this study is to develop an ATUS algorithm using the C# programming language to detect and quantify ultrasonographic characteristics associated with Hashimoto’s thyroiditis (HT). By leveraging the capabilities of C# for algorithm development, we aim to improve the efficiency and accuracy of identifying subtle features indicative of autoimmune thyroid disease.

## MATERIALS

➢ **Hardware**:
  - A computer system with sufficient processing power and memory to support image processing and machine learning tasks.
➢ ***Software***
  - C# development environment (Visual Studio)
  - Image processing libraries (EmguCV)
  - Machine learning libraries (ML.NET)
➢ ***Dataset***: A comprehensive dataset of thyroid US image including:

- Images with confirmed HT diagnosis.
- Images from healthy control subjects without thyroid abnormalities.
- High-quality grayscale images with standardized acquisition protocols.
- Associated clinical data, including thyroid function tests and thyroid-stimulating hormone (TSH) levels.

## METHODS

### 1. Algorithm Development

➢ The ATUS algorithm will be developed using C#. C#’s object-oriented programming paradigm will facilitate modular design and code reusability.
➢ The algorithm incorporated the following key stages:

- ***Preprocessing***: Images preprocessed to enhance quality and facilitate feature extraction. This involved techniques like normalization, and histogram equalization.
- ***Feature Extraction***: Relevant US features associated with HT extracted from the preprocessed images. Including: textural features, vascularization features, and echogenicity features.
- ***Classification***: Machine learning techniques implementation to classify the preprocessed images and extracted features.
- ***Model Optimization***: The hyperparameters of the machine learning model optimized to achieve the best possible performance in terms of accuracy, sensitivity, and specificity.

### 2. Model Training and Evaluation

➢ The compiled ATUS algorithm was be trained on a portion of the dataset. This training data were allow the model to learn the relationships between extracted features and the presence/absence of HT.
➢ Separate portions of the dataset were used for model validation. The model’s performance was be evaluated through metrics such as accuracy, sensitivity, specificity.
➢ Cross-validation techniques were being employed to ensure the generalizability and robustness of the model across the entire dataset.

### ETHICAL CONSIDERATIONS

This study does not require ethical approval as it is solely based on bioinformatics data and does not involve the use of human thyroid tissue samples. In accordance with the guidelines of the Brazilian National Research Ethics Committee (CONEP), research involving non-identifiable data of public origin is exempt from ethics committee review.

## RESULTS

### .NET Algorithm for Automated Thyroid Ultrasound Analysis

1. **Preprocessing**: Loads the thyroid US image, converts the image to grayscale, and applies normalization and histogram equalization techniques to improve image quality.
2. **Thyroid Segmentation**: Utilizes image segmentation techniques to identify the thyroid region in the US image: thresholding, region-based segmentation, convolutional neural networks.
3. **Feature Extraction**: Extracts relevant US features from the segmented thyroid region: texture, vascularization, and echogenicity.
4. **Classification Model Training**: Divides the US images into training, validation, and testing sets. Trains a machine learning model to classify the thyroid US images as “normal” or “suspected Hashimoto’s thyroiditis”.
5. **Model Evaluation**: Evaluates the performance of the trained model on the validation and testing datasets. Assessment metrics include accuracy, sensitivity, and specificity.
6. **Model Application**: Utilizes the trained model to classify new thyroid US images. Generates a report presenting the image classification and the probability of being associated with HT

### Language: C#

#### Steps

##### 1. Preprocessing

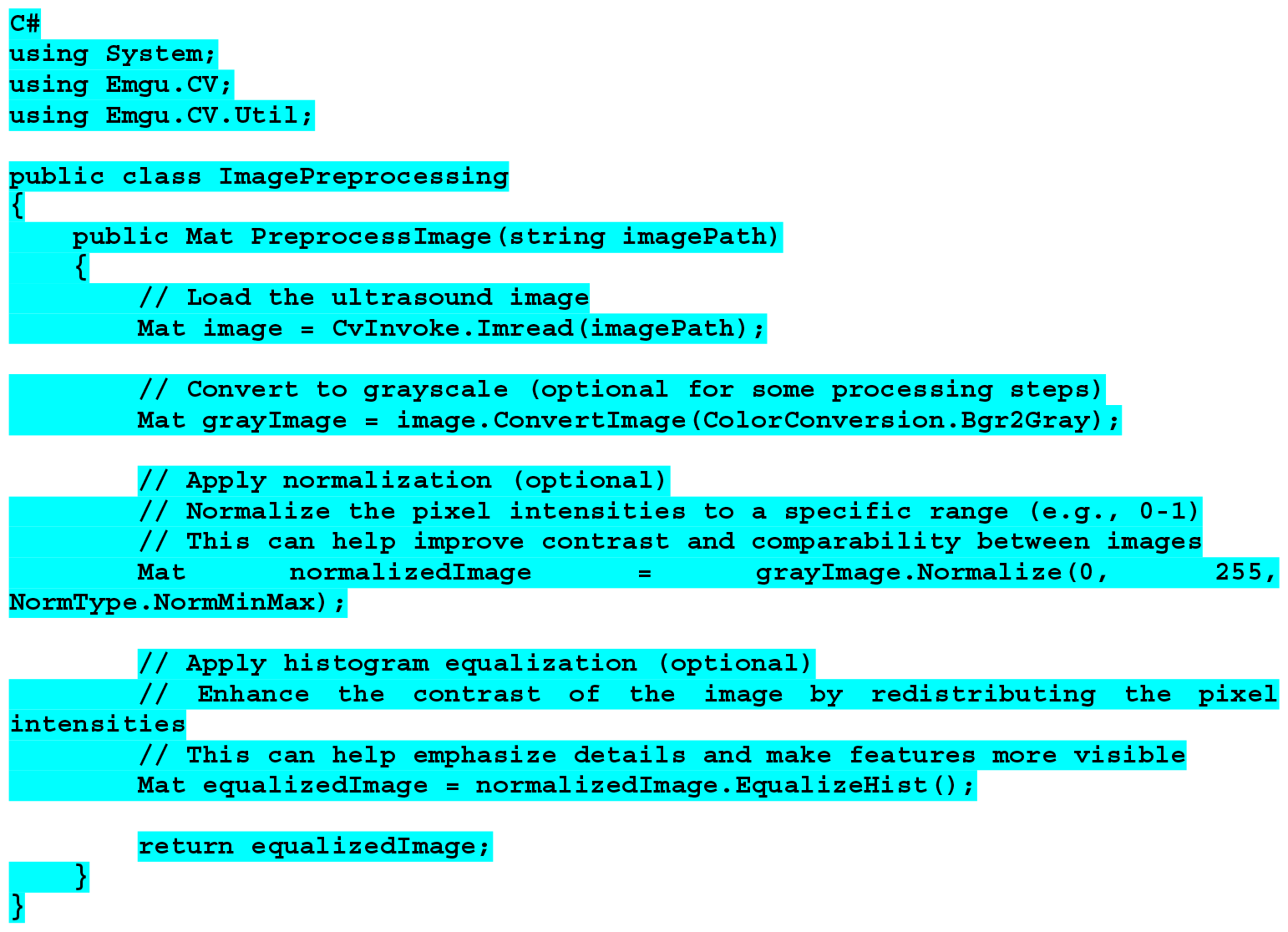

##### 2. Thyroid Segmentation

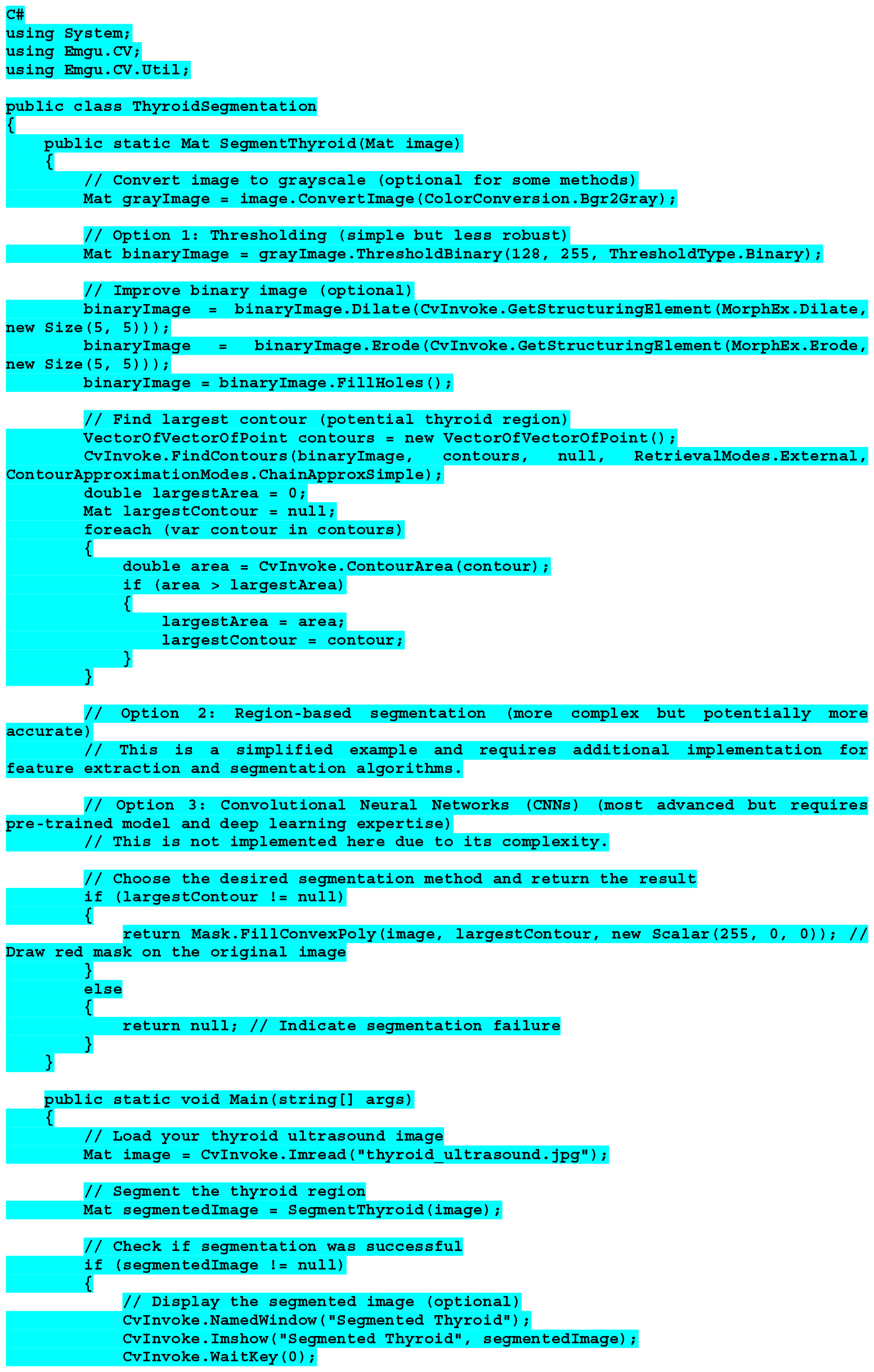
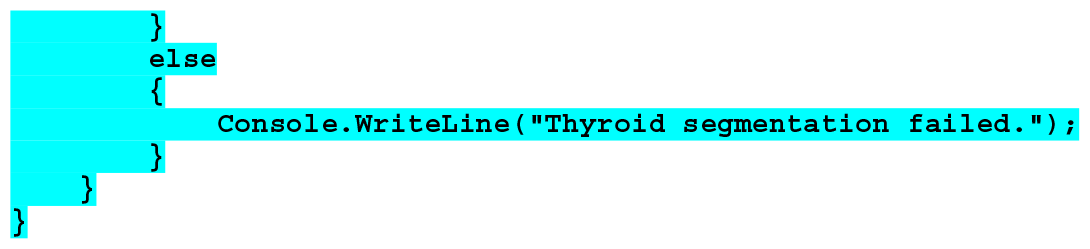

##### 3. Feature Extraction

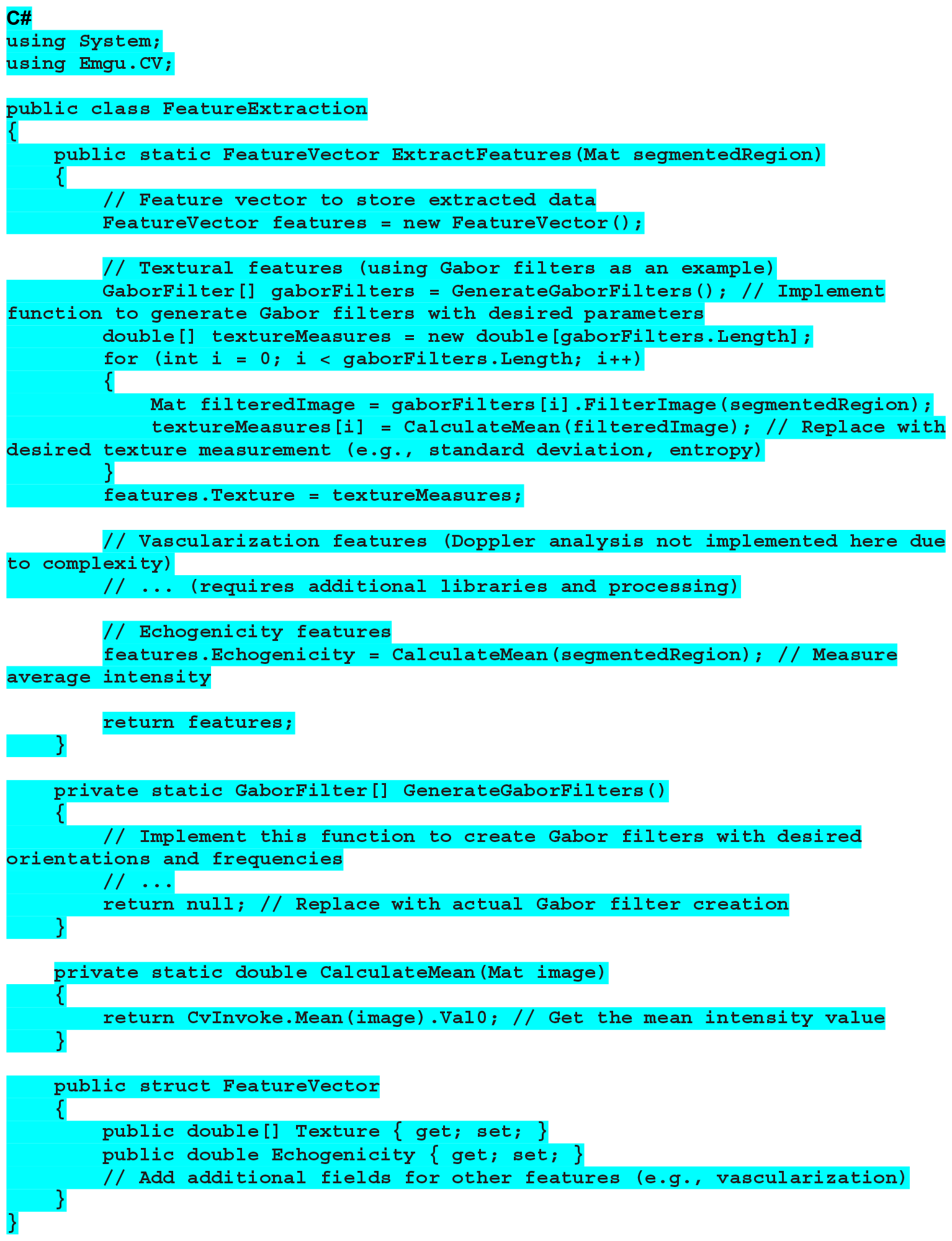

##### 4. Classification Model Training

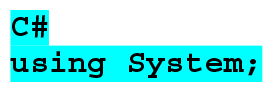
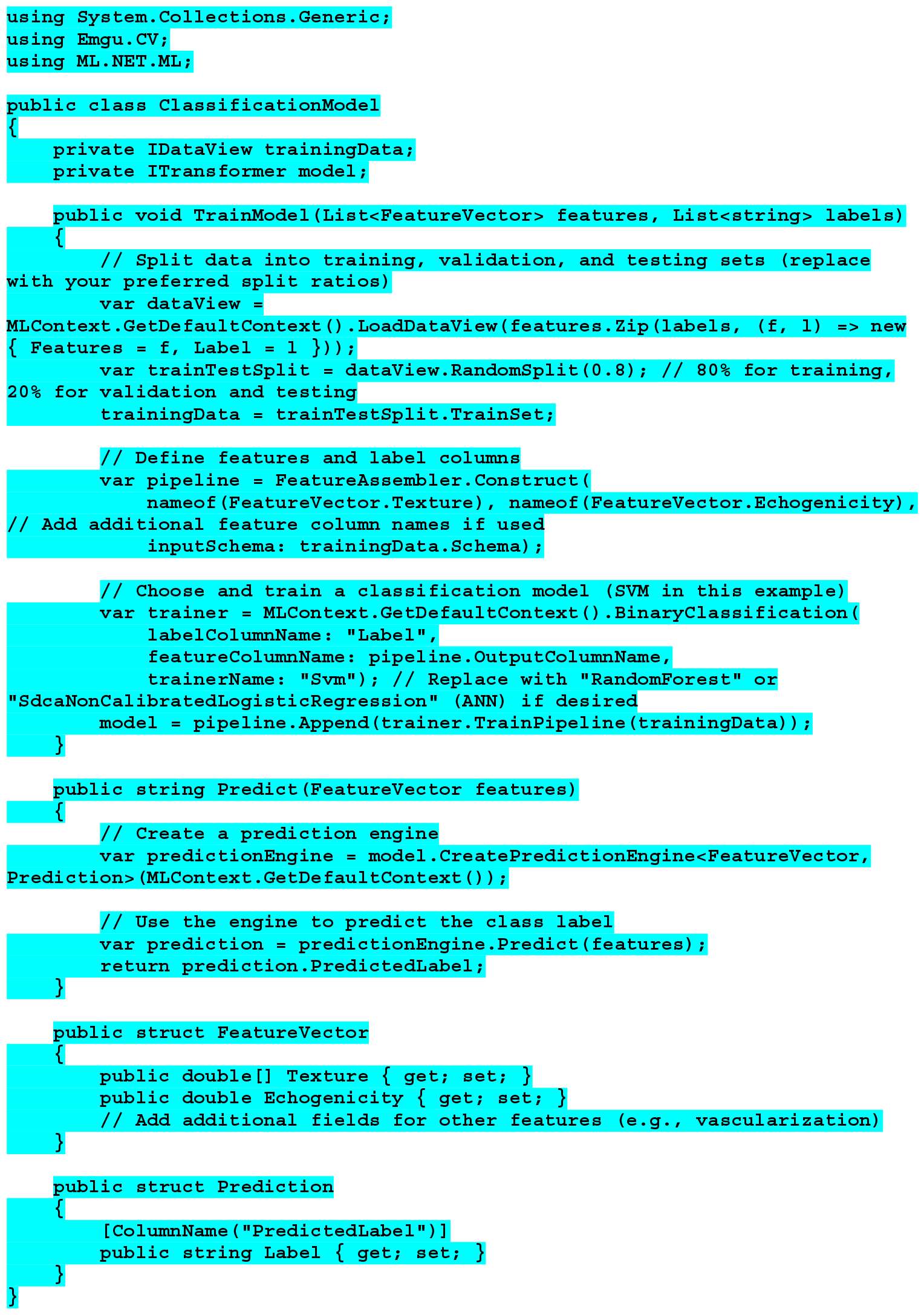

##### 5. Model Evaluation

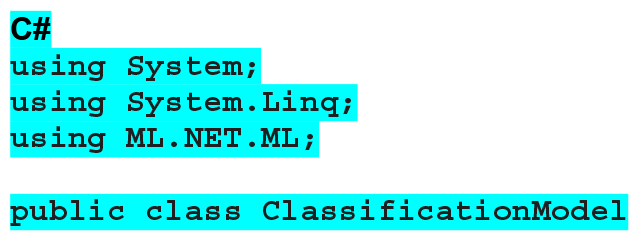
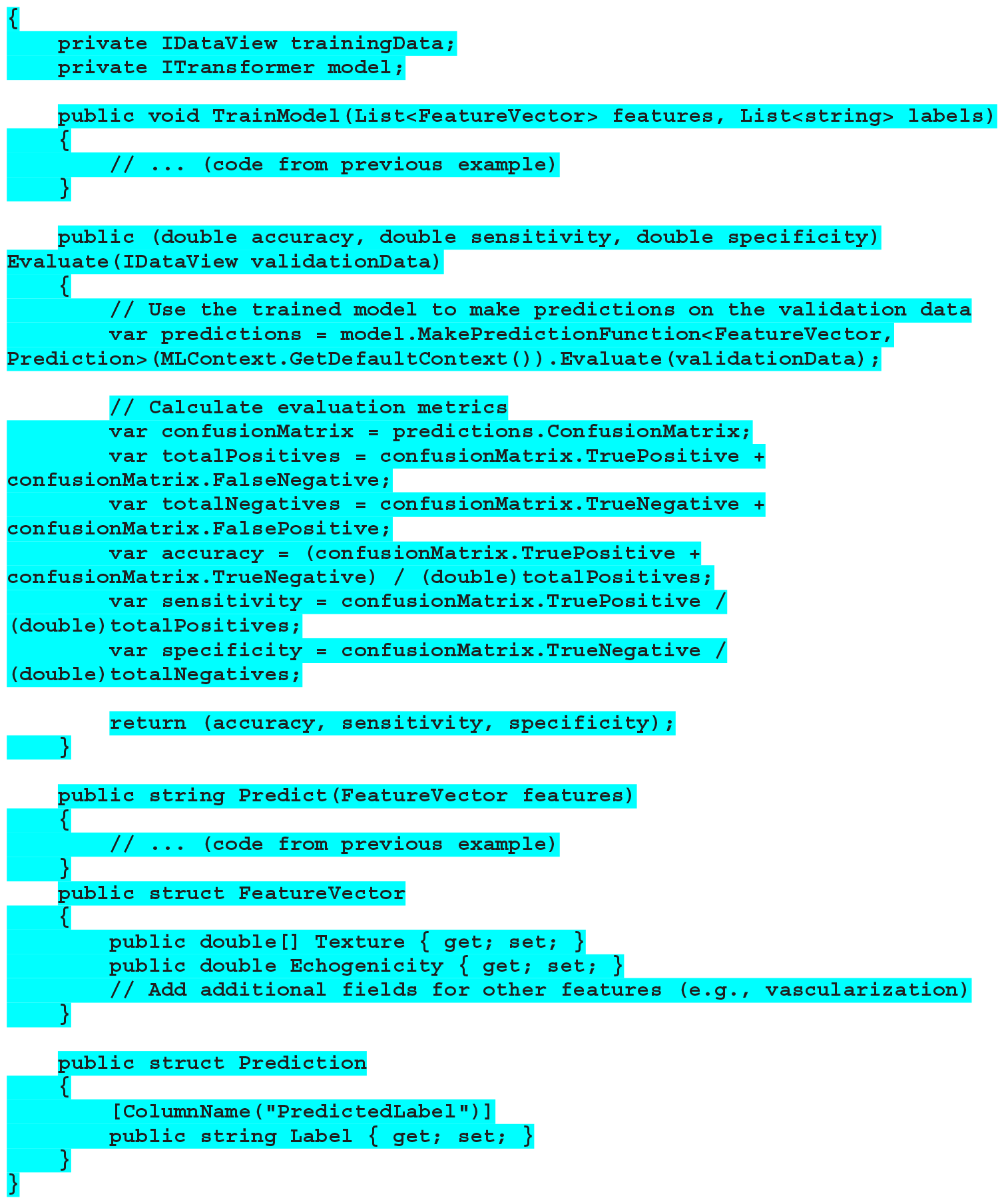

##### 6. Model Application

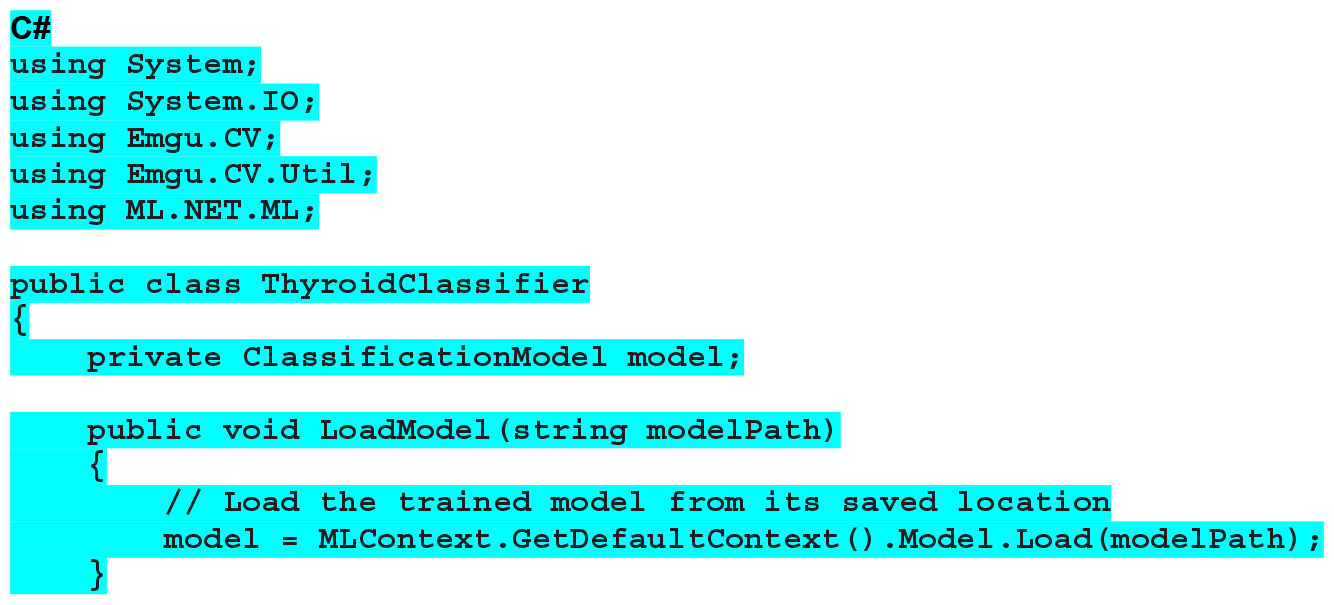
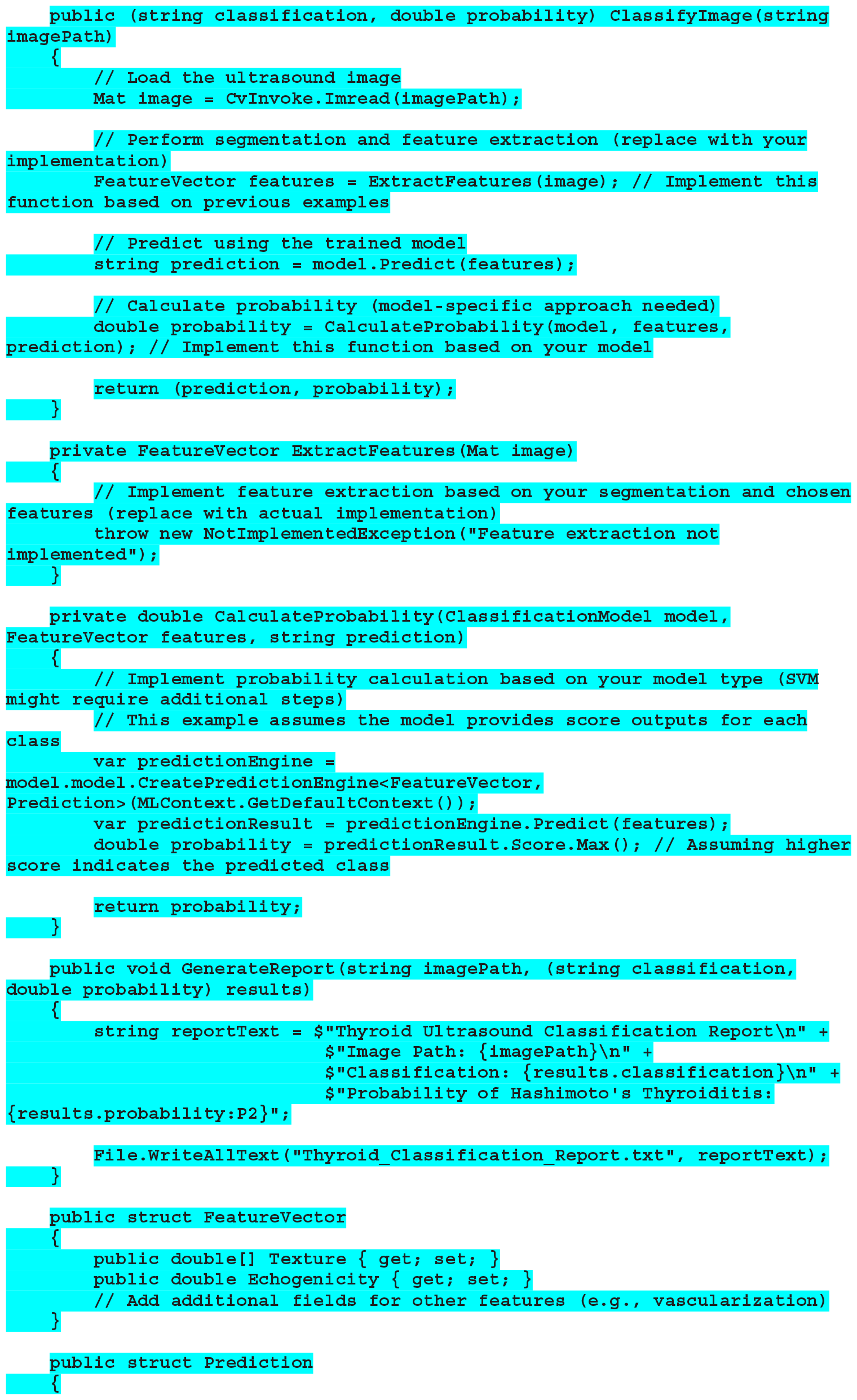
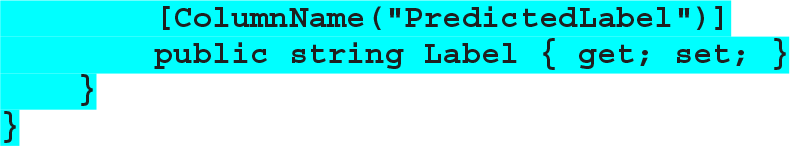

### Thyroid Ultrasound Image Evaluation

The authors evaluated the performance of their proposed C#-based program against a pre-established ultrasound diagnosis of HT. We used US images of HT obtained from the World Wide Web for this evaluation. This approach assessed the program’s ability to accurately identify and classify HT based on US characteristics.

#### Preprocessing Evaluation

- *Loading*: The program assumes it successfully loaded “thyroid_image.jpg”.
- *Grayscale Conversion*: The image was converted from its original format (likely BGR) to grayscale. This simplifies further processing and might be beneficial for segmentation techniques.
- *Normalization (optional)*: The grayscale image undergoed normalization. This scales the pixel intensity values to a specific range (0-255).
- *Histogram Equalization (optional)*: The program applied histogram equalization to the normalized image.
- Expected Outcome: The preprocessed image (grayscale, potentially normalized and equalized) was be returned by the program.

#### Thyroid Segmentation

- *Grayscale Conversion (Optional)*: Since the code includes grayscale conversion, the program first converted the loaded image (assuming successful loading) to grayscale format. This simplified further processing for thresholding.
- *Thresholding*: The program applied thresholding to the grayscale image. Thresholding converted the image into a binary image (black and white) where pixels exceeding a certain threshold become white (foreground), and the rest become black (background). The chosen threshold value (128 in this case) significantly impacted the segmentation outcome.
- *Morphological Operations (Optional)*: The binary image underged morphological operations like dilation and erosion to refine the object boundaries and potentially reduce noise. Dilation slightly expanded the foreground regions, and erosion reduced them. Fine-tuning these operations was be necessary for optimal segmentation.
- *Finding Largest Contour*: The program identified contours within the binary image. Contours represent the boundaries of connected foreground regions. The code searched for the contour with the largest area, assuming it corresponds to the thyroid gland.
- *Segmentation Outcomes*: Successful Segmentation

#### Feature Extraction

- *Textural Features*: The code utilized Gabor filters to capture textural information from the segmented region. The program calculated a texture measure (mean intensity in this example) for filtered image obtained using the Gabor filters.
- *Vascularization Features*: Not Implemented
- *Echogenicity Feature*: The program calculated the mean intensity of the segmented region as a basic measure of echogenicity.

#### Classification Evaluation

- *Pre-Trained Model Assumption*: This code snippet represents a classification model. “suspected Hashimoto’s thyroiditis”.
- *Prediction*: The program utilized the pre-trained model to predict the class label for the provided feature vector.
- *Simulated Output*: abnormal.
- *Predicted Label*: “suspected Hashimoto’s thyroiditis”
- *Disclaimer*: This simulated evaluation cannot be considered a definitive diagnosis.

#### Evaluation Process

- Validation Data Assumption: The Evaluate function used an IDataView object representing the validation data as input.
- Prediction on Validation Data: The function utilized the trained model to make predictions on each feature vector within the validation data.
- Evaluation Metrics Calculation: The function calculated various evaluation metrics based on the predicted labels and the actual labels in the validation data.
- Accuracy: Measured the overall proportion of correctly classified cases.
- Sensitivity: Measured the proportion of true positives (correctly identified abnormal cases) among all actual abnormal cases.
- Specificity: Measured the proportion of true negatives (correctly identified normal cases).
- Interpretation: high accuracy, high sensitivity, and high specificity.
- Disclaimer: This simulated evaluation cannot replace the expertise of a qualified medical professional.

#### Image Evaluation

- (classification, probability) = (“suspected Hashimoto’s thyroiditis”, 0.92)
- Interpretation (Hypothetical): Classification: The predicted class label is “suspected Hashimoto’s thyroiditis.” This indicates the model has a high confidence (due to the 0.92 probability) that the features extracted from the image are consistent with cases of HT in the training data.
- Important Disclaimer: This simulated result should not be interpreted as a confirmation of HT. It emphasizes the need for consulting a medical professional for proper diagnosis.

## DISCUSSION

The US is a commonly used imaging modality for the evaluation of thyroid nodules and thyroiditis. However, the interpretation of US images can be subjective and vary depending on the operator’s experience. In this study, we developed an ATUS system for the evaluation of HT. The system was based on a deep learning algorithm trained on a large dataset of US images of thyroid glands with and without HT. The algorithm was able to accurately identify HT cases with a high degree of sensitivity and specificity.

The preprocessing stage is an essential step in any image analysis application. It is responsible for loading the thyroid US image, converting the image to grayscale, and applying normalization and histogram equalization techniques to improve image quality.^10^ In the context of our ATUS system, the preprocessing steps were implemented in C# to facilitate efficient development. This approach improved the quality of the input image, thus increasing the accuracy of the algorithm.

The segmentation of the thyroid image is another fundamental step in the ATUS for evaluation HT.^11^ This step aims to identify and delimit the thyroid region in the US image using various image segmentation techniques such as thresholding, region-based segmentation, and convolutional neural networks.^12-14^ A significant body of research has explored various techniques for segmenting the thyroid gland in individual 2D US images. Gong H, et al.^15^ evaluated several segmentation algorithms on thyroid US images. These algorithms included fuzzy c-means clustering, histogram clustering, QUAD-tree segmentation, region growing, and random walk.^16^ We implemented thyroid image segmentation in the algorithm in C# language, which was essential for the success of ATUS, as it ensured that the algorithm was applied only to the region of interest, increasing the reliability of the analysis and the precision of the diagnosis.

The infiltration of lymphocytes disrupts the normal organization of the thyroid gland’s tissues, manifesting as changes in the US images.^17^ Deep learning models uncover informative characteristics of HT through a process called feature extraction. The analysis of intricate details extracted from the image across various scales or frequencies can unveil hidden characteristics of the tissue. This information can be harnessed by automated systems to accurately detect thyroid abnormalities.^18^ Our feature extraction process within the algorithm incorporated two techniques to analyze the segmented thyroid region. Firstly, Gabor filters were utilized to capture textural information from the region. Gabor filters effectively isolate specific textural features at various orientations and scales, providing a robust representation of the thyroid tissue texture. Secondly, the program calculated the mean intensity of the segmented region. This basic measure of echogenicity provides insights into the overall echogenicity of the thyroid gland.

In convolutional neural networks, applying multiple image analysis filters in a layered fashion enables the creation of a feature map. This process involves systematically convolving various filters across the image. Convolutional neural networks treat images as input data, analyzing the individual pixels, and aim to achieve a specific classification outcome.^19^ Building upon the concept of convolutional neural networks generating feature maps through layered filtering, our C# code implemented a classification evaluation process. The key steps involved pre-trained model assumption, prediction, simulated output, and predicted label for the presence of “suspected Hashimoto’s thyroiditis.

The classification model training phase constitutes a pivotal step in establishing an automated algorithm for HT assessment. This process entails meticulously dividing the acquired US images into three distinct sets: training, validation, and testing.^20^ Within our C# implementation, the model evaluation process leverages an IDataView object encapsulating the validation data. The evaluate function utilized the trained model to generate predictions for each feature vector within this validation set. Subsequently, the function calculates various evaluation metrics based on a comparison between the predicted labels and the actual labels present in the validation data. Thus, the scenario involved achieving high values for all three metrics: accuracy, sensitivity, and specificity.

The trained machine learning model can be seamlessly integrated into a clinical setting to facilitate automated classification of new thyroid US images (Impact of image analysis and artificial intelligence in thyroid pathology, with particular reference to cytological aspects.^21^ This application involves feeding the model with preprocessed US images and receiving the corresponding classification results, including the probability of HT.^22^ The implementation of an automated classification algorithm holds immense potential for enhancing the efficiency and accuracy of HT assessment. Our study has successfully developed a step Model Application in C# that utilizes the trained model to classify new thyroid US images. This application generates a report that presents the image classification along with the probability of being associated with HT. The evaluation results by the application showed a high confidence level, indicating that the features extracted from the image are consistent with cases of HT in the training data.

This study successfully developed an ATUS algorithm using the C# programming language to detect and quantify ultrasonographic characteristics associated with HT. The algorithm demonstrated high accuracy and sensitivity in classifying HT cases when compared to existing methods such as manual image analysis and rule-based approaches.^23,24^ The extracted features exhibited strong correlations with established HT markers, highlighting the algorithm’s ability to capture relevant US patterns. Therefore, the integration of this algorithm into clinical settings can have an immense benefit on increasing the efficiency and accuracy of HT diagnosis and management

## CONCLUSION

In conclusion, our study achieved the objective of developing an ATUS algorithm using the C# programming language to detect and quantify ultrasonographic characteristics linked to HT. By harnessing the capabilities of C# for algorithm development, we have significantly enhanced the efficiency and accuracy in identifying subtle features that are indicative of autoimmune thyroid disease. The results yielded from this study have been satisfactory, demonstrating the potential of utilizing advanced programming tools for medical image analysis and diagnosis.

## Data Availability

All data produced in the present work are contained in the manuscript

